# Surgical Guidance of Head and Neck Squamous Cell Carcinoma Enabled by a c-MET-Targeting Fluorescent Probe cMBP-ICG

**DOI:** 10.1101/2022.06.02.22272990

**Authors:** Jingbo Wang, Siyi Li, Kun Wang, Ling Zhu, Lin Yang, Yunjing Zhu, Zhen Zhang, Longwei Hu, Yuan Yuan, Qi Fan, Jiliang Ren, Gongxin Yang, Weilong Ding, Xiaoyu Zhou, Ruimin Huang, Junqi Cui, Chunye Zhang, Ying Yuan, Jie Tian, Xiaofeng Tao

**Affiliations:** Department of Radiology, Shanghai Ninth People’s Hospital, Shanghai Jiao Tong University, Shanghai, 200011, China; Department of Oral and Maxillofacial-Head and Neck Oncology, Ninth People’s Hospital, School of Medicine, Shanghai Jiao Tong University, Shanghai, 200011, China; CAS Key Laboratory of Molecular Imaging, Beijing Key Laboratory of Molecular Imaging, The State Key Laboratory of Management and Control for Complex Systems, Institute of Automation, Chinese Academy of Sciences, Beijing, 100000, China; Molecular Imaging Research Center, Shanghai Institute of Materia Medica, Shanghai, 200011, China; Department of Pathology, Shanghai Ninth People’s Hospital, Shanghai Jiao Tong University, Shanghai, 200011, China

**Keywords:** near-infrared fluorescence dye, topical application, c-MET, head and neck squamous cell carcinoma

## Abstract

**Importance:** Head and neck squamous cell carcinoma (HNSCC) is a highly aggressive malignancy which is associated with subtle local invasion and metastasis. Near-infrared (NIR) fluorescence-guided surgical precise resection of the tumor margin and potential metastasis has been appealing.

**Objective:** To determine whether a peptide-based NIR fluorescence dye, which can be administered via topical application and target tumor tissue via c-MET binding, can improve discrimination of HNSCC from normal tissue.

**Design:** A nonrandomized controlled trial was conducted between March 2021 and February 2022 at Shanghai Ninth Peoples’ Hospital, China. Study participants were blinded for subjective measurements.

**Setting:** Exploratory, single-center, open-label, dose-escalation

**Participants:** Consecutive Asian individuals with clinically suspicious, primary or recurrent HNSCC, who were scheduled for standard-of-care surgery with curative intent, were enrolled.

**Interventions:** Enrolled patients underwent in vivo fluorescence imaging before surgery, and ex vivo fluorescence imaging was performed on intraoperative specimens using 2.5 μM or 5.0 μM topical applicated fluorescent probes.

**Main Outcomes and Measures:** Primary analysis compared mean fluorescence intensity (MFI, total fluorescence signal divided by the total number of pixels within the region of interest) of tumor and peritumoral normal tissue. Possible tumor-to-background ratios (TBR, ratio of MFI of tumor over peritumoral normal tissues) may range from 2 to 4.

**Results:** Ten patients (1 female [10%], age = 59.4 ± 12.6 years old) were enrolled. Ex vivo imaging with 5.0 μM fluorescence prob demonstrated TBR of 2.71 ± 0.7, with 2.5 μM fluorescence prob demonstrated TBR of 3.11 ±1.2. To discriminate tumor from normal tissue with MFI, the area under the curve (AUC) was 0.88 (95% CI-0.88 to 1.0) for the 5.0 μM group, and 0.65 (95% CI 0.53 to 0.83) for the 2.5 μM group.

**Conclusions and Relevance:** This trial demonstrates that topical application of tumor targeting fluorescence probe can differentiate tumor and outlining tumor margins. Further research is currently underway to estimate whether this probe is potent in guiding biopsy and surgical procedure, and assisting long-term follow-up postoperatively.

**Trial Registration:** Chinese Clinical Trial Registry Identifier: ChiCTR2200058058

## 1. Introduction

Head and neck squamous cell carcinoma (HNSCC) accounts for 5% of all systemic malignant tumors, with more than half a million new cases each year.^1,2^ Despite recent advances in medical development, the survival rate of HNSCC patients has not been effectively improved.^3^ Appropriate surgical resection range can preserve the oral and facial function of patients as much as possible and also reduce the local recurrence rate. However, due to the characteristics of invasion and heterogeneity of HNSCC, there is still no recognized standard for the resection range of this tumor.^4^

Over the past decade, near-infrared (NIR) fluorescence-guided surgery has emerged as a promising technique for head and neck cancers using tumor-targeting fluorescent agents (e.g., panitumumab-IRDye800CW, cetuximab-IRDye800CW^5,6^). For local small-distance metastases, extracellular matrix (ECM) may prevent the penetration of intravenously administered macromolecular agents ^7^, which may lead to different mean fluorescence intensity (MFI) and tumor background ratio (TBR), thereby causing a higher miss rate for small lesions. Therefore, the intravenous delivery of NIR fluorescence dye may not be suitable for shallow, invasive, and heterogenous tumors, such as HNSCC.

Here, we propose that topically applied, NIR fluorescently labeled, simply synthesized homing-peptide–based, head and neck tumor–targeting contrast agent, cMBP-ICG, can leverage the deficiency of intravenously administrated antibody-based agents discussed above. cMBP is a c-MET–targeting peptide that has been developed for tumor imaging and gene delivery^8-10^. The mouth-washing topical application of the targeting imaging agents may be an optimal approach given that mouthwash covers the entire oral mucosa and directly screens out suspicious small cancer foci without restriction from the other tissues. Being applied topically, the imaging time of NIR fluorescence probes would be significantly reduced (within minutes)^11^. Peptide-based targeting agents’ fast renal clearance^12^ is not seen as a drawback, but rather a benefit from the perspective of medication safety.

In this study, we aimed to determine the safety and feasibility of cMBP-ICG topical application for real-time NIR fluorescence imaging. To our knowledge, this study describes the first clinical application of cMBP-ICG in patients with HNSCC for preoperative *in vivo* and intraoperative *ex vivo* detection of primary tumor and metastasis.

## 2. Methods

### 2.1. Clinical Study Design and Participants

An exploratory, single-center, open-label, dose-escalation study was approved by the institutional review board of Chinese Clinical Trial Registry (ChiCTR2200058058). Written informed consent was obtained from all patients. Between March 2021 and February 2022, a total of 10 patients with clinically suspicious, biopsy-proven, primary or recurrent HNSCC were enrolled (**Table 1**), and were scheduled to undergo standard-of-care surgery with curative intent at Shanghai Ninth Peoples’ Hospital. All of the enrolled patients were aged 18 years or older, with a life expectancy of more than 12 weeks. Exclusion criteria and the entire flowchart are shown in **Figure 1**. All of the enrolled patients underwent preoperative imaging (CT and/or MRI). TNM stage was determined in accordance with the American Joint Committee on Cancer (AJCC) eighth edition criteria^13^.

**Table 1.**
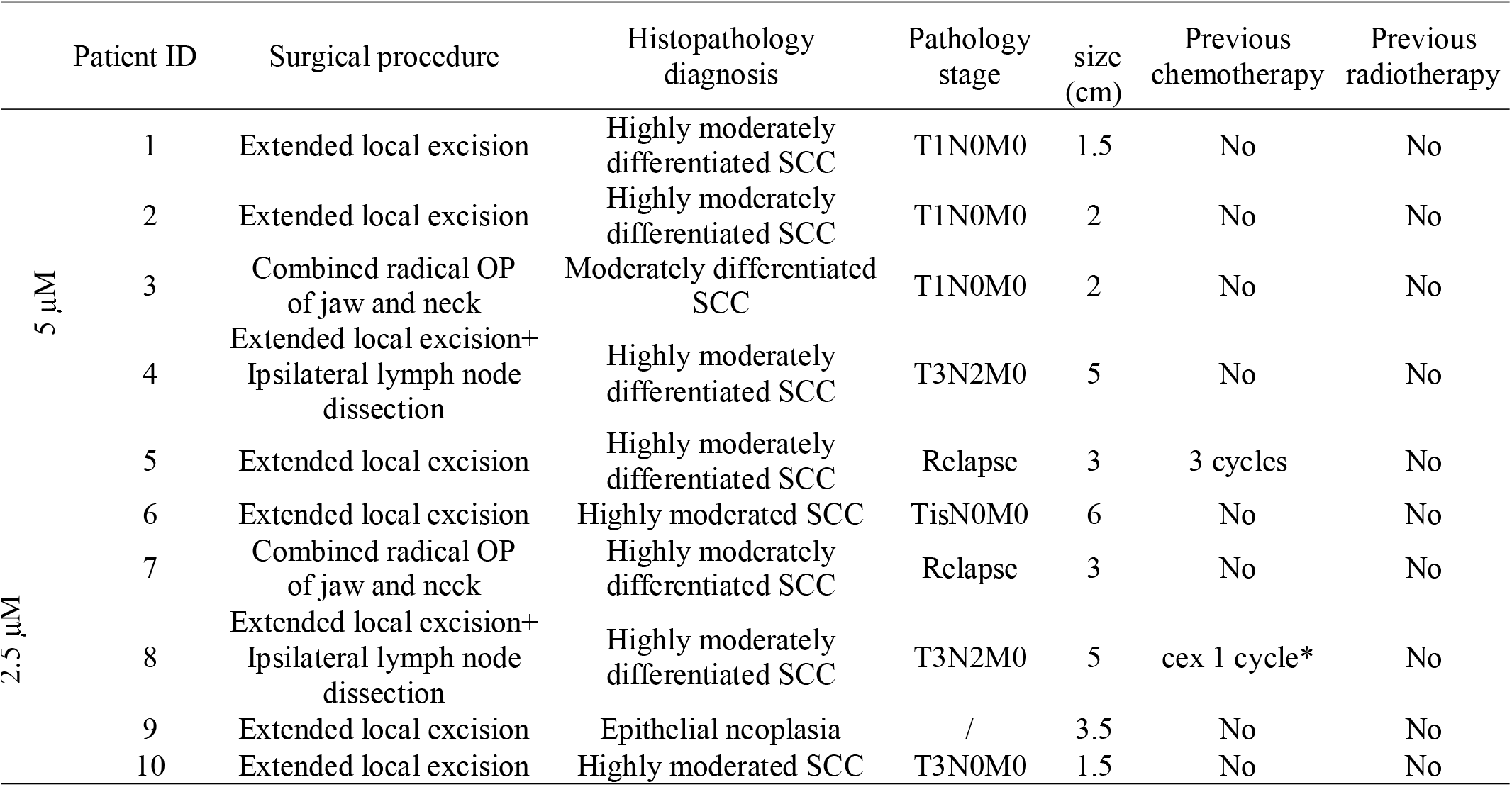
Demographic and clinical information for 10 cases applied in clinical trials

|  | Patient ID | Surgical procedure | Histopathology diagnosis | Pathology stage | size (cm) | Previous chemotherapy | Previous radiotherapy |
| --- | --- | --- | --- | --- | --- | --- | --- |
| 5 μM | 1 | Extended local excision | Highly moderately differentiated SCC | T1N0M0 | 1.5 | No | No |
|  | 2 | Extended local excision | Highly moderately differentiated SCC | T1N0M0 | 2 | No | No |
|  | 3 | Combined radical OP of jaw and neck | Moderately differentiated SCC | T1N0M0 | 2 | No | No |
|  | 4 | Extended local excision+ Ipsilateral lymph node dissection | Highly moderately differentiated SCC | T3N2M0 | 5 | No | No |
|  | 5 | Extended local excision | Highly moderately differentiated SCC | Relapse | 3 | 3 cycles | No |
| 2.5 μM | 6 | Extended local excision | Highly moderated SCC | TisN0M0 | 6 | No | No |
|  | 7 | Combined radical OP of jaw and neck | Highly moderately differentiated SCC | Relapse | 3 | No | No |
|  | 8 | Extended local excision+ Ipsilateral lymph node dissection | Highly moderately differentiated SCC | T3N2M0 | 5 | cex 1 cycle* | No |
|  | 9 | Extended local excision | Epithelial neoplasia | / | 3.5 | No | No |
|  | 10 | Extended local excision | Highly moderated SCC | T3N0M0 | 1.5 | No | No |

**Figure 1.**
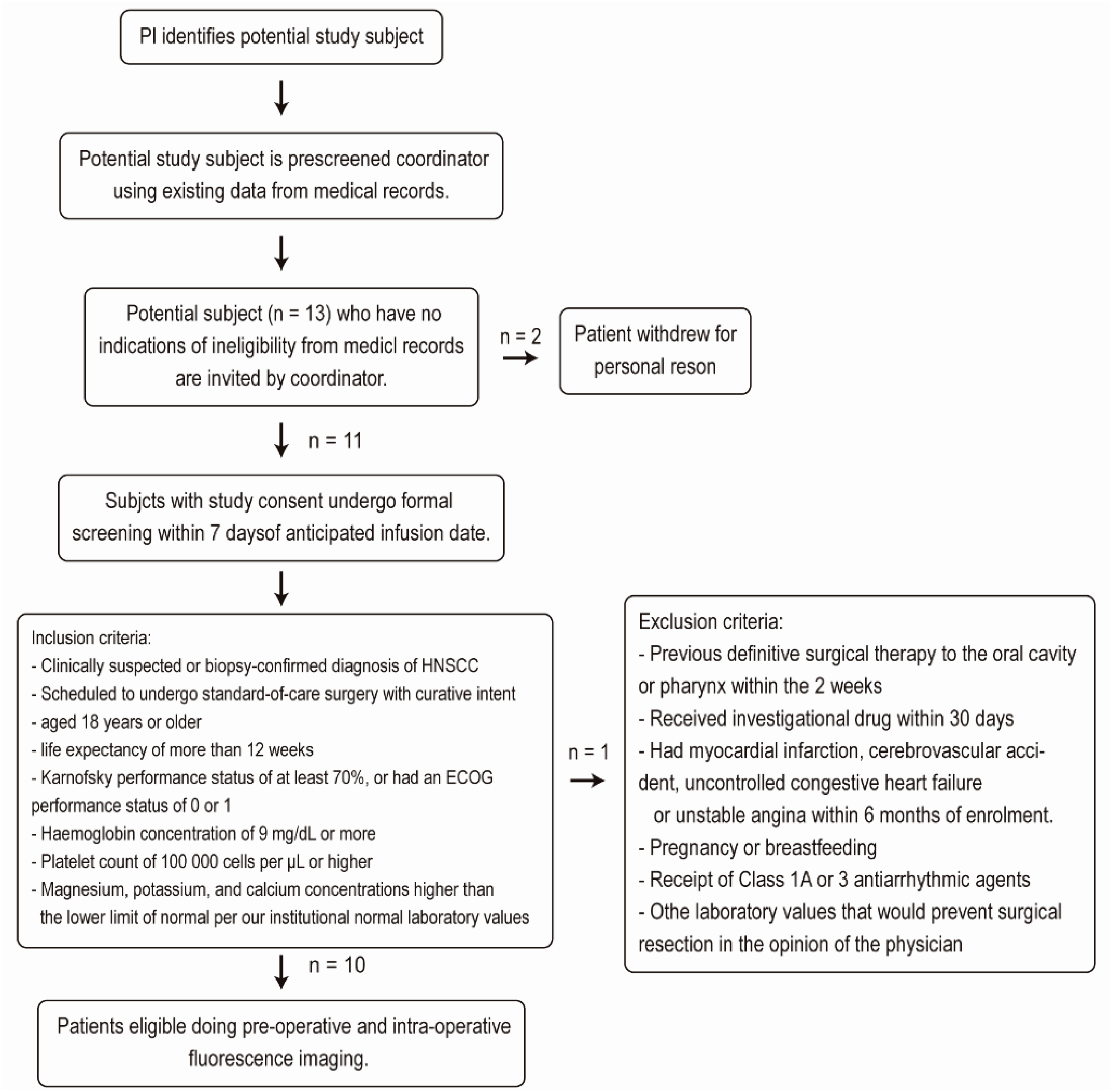
Flow Diagram Representing Recruitment Process

### 2.2. Procedures of the Clinical Trial

Lyophilized powder of cMBP-ICG (1.0 mg, 3.6×10^−4^ mmol) was dissolved in ultrapure water (145.7 mL or 72.8 mL) to a concentration of 2.5 μM or 5.0 μM. In order of enrollment, the first five cases used a 5.0 μM concentration for gargling or ex vivo specimen surface smearing, and the next five cases used a 2.5 μM concentration. For preoperative *in vivo* cMBP-ICG application, the patients gargled 25 mL of cMBP-ICG solution for 30 s, and then spit it out (twice). The patients gargled 30 mL of ultrapure water for 30 s and spit it out (twice). A preclinical fluorescence laryngoscope imaging platform (DPM-III-01, Zhuhai Dipu Medical Technology Co., Ltd.) was used for real-time fluorescence imaging. For imaging investigations, videos were acquired before topical application, after topical application pre-wash, and after topical application post-wash using a fluorescence laryngoscope. The camera lens was aimed at ROI and manually focused. The same instrument settings were used for all of the imaging procedures (exposure time, 30 ms; excitation power, 20%; gain: 3 dB; at 60 frames per second). The distance between the lens and surface of ROI was kept at around 10 mm^14^. The videos were acquired by slowly scanning the tumor area and surrounding normal mucosa (non-tumor region).

The patients then underwent surgical operation on the next day. For intraoperative *ex vivo* back-table fluorescence-guided imaging (FGI), the resected tumor specimens were collected directly after excision, and the fresh (non-fixed) samples were incubated with cMBP-ICG at either of the abovementioned concentrations. Then, the samples were rinsed for 1 min with PBS to clear unbound cMBP-ICG (twice), followed by direct NIR fluorescence imaging. Because the fluorescent probe is coated on the surface of resected tumor specimens, the fluorescence images cannot be directly compared with the cross section. Small tissues were cut off from specimens’ high/low fluorescence intensity surface respectively. The primary resecting surgeon then predicted whether these tissues contained malignancy according to their visual features and tactile textures. The surgeon’s predicted outcome was compared with that of fluorescence imaging. The distance between the lens and the tissue surface was kept at approximately 20 cm for ex vivo imaging.

Histopathologic processing of the surgical specimens was performed by a board-certified head and neck pathologist. Intraoperatively, frozen-section staining was routinely performed with hematoxylin and eosin with real-time guidance from NIR fluorescence imaging. The remaining tumor tissue was embedded in paraffin, and 5-μm-thick sections were obtained. Two neighboring sections were stained with H&E and IHC for c-MET. A board-certified pathologist, who was masked to NIR fluorescence results, evaluated the H&E slides microscopically and outlined the tumor areas. In the analysis of ex vivo tissue, the pathology diagnosis was used as the gold standard for tumor detection.

Adverse events were reported on the basis of federal regulations^15^ and the National Cancer Institute Common Terminology Criteria for Adverse Events (v5.0). The workflow of the clinical trial is shown in **Figure 2**.

**Figure 2.**
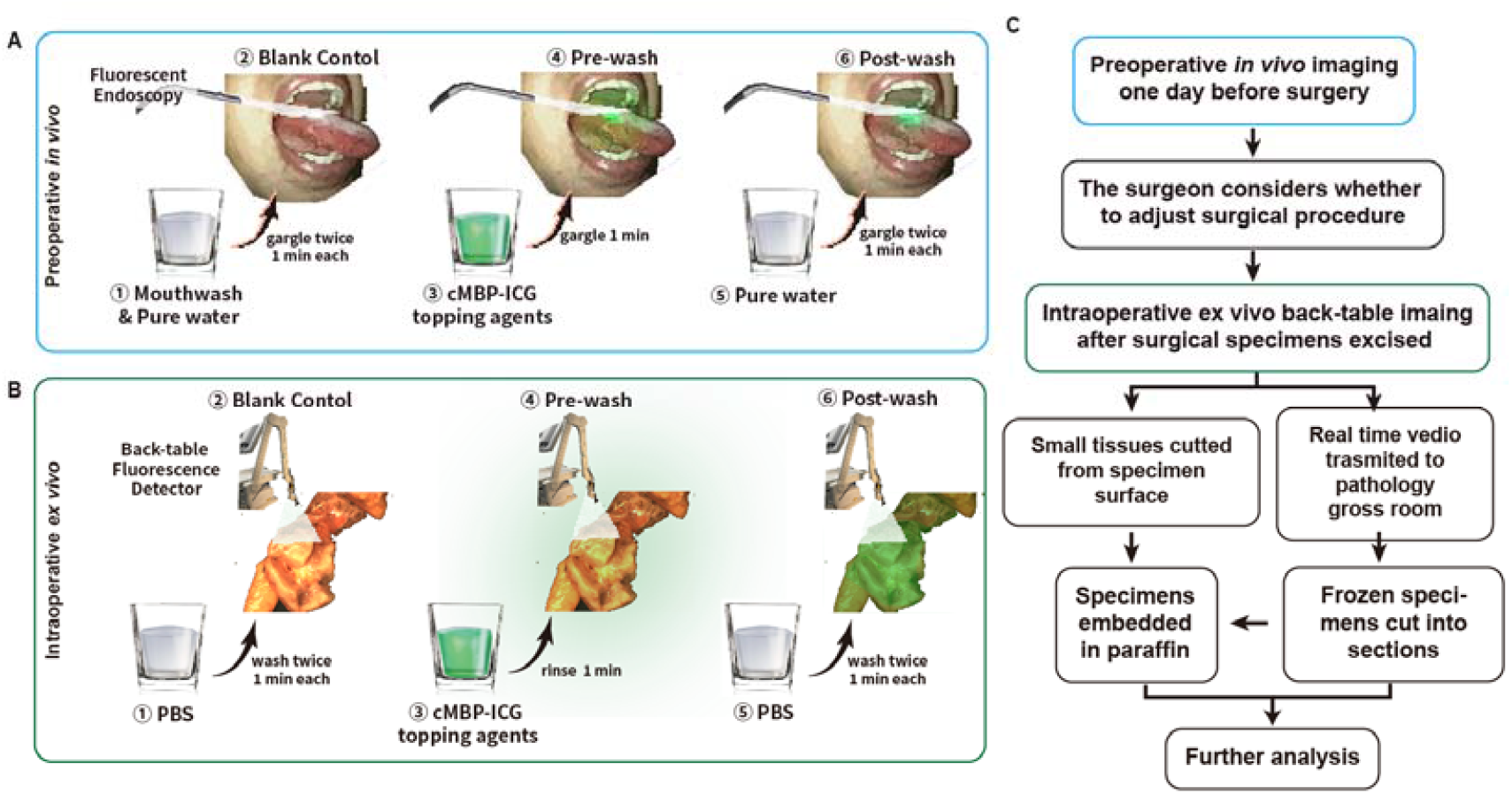
Workflow of fluorescence imaging-based HNSCC delineation. **A** After careful mouth washing (step 1), patients (n = 10) with clinically suspicious or biopsy-proven head and neck squamous cell carcinoma (HNSCC) were examined using a Digital Precision Medicine (DPM, Beijing, China) imaging device H2000, with an endoscopic camera and ICG-optimized LED–filter system. The tumor area and the surrounding margins were imaged as a blank control (step 2). After gargling a solution of cMBP-ICG (step 3) at different concentrations (2.5 μM, 5 μM, each dose n = 5) for 1 min, a pre-wash imaging step was performed (step 4). Then, the patients gargled ultrapure water for 1 min as a clearing solution (step 5), followed by cMBP-ICG administration post-wash imaging (step 6). **B** Immediately after resection, tissue specimens, including margin samples, were imaged ex vivo in a vertical near-infrared imaging device Z2000 (DPM, Beijing, China) following the same protocol described above. **C** Preoperative images were taken into consideration during surgical procedure plan. Intraoperative back-table videos were transmitted in real time to pathology gross room as frozen section’s references. Small tissues were cut off from specimens’ high/low fluorescence intensity surface respectively for further analysis.

### 2.2. Data Analysis and Statistical Analysis

For the image analysis, ImageJ ^16^ software was used. The tumor-to-background ratio (TBR) was determined as the ratio of fluorescence intensity of tumor region of interest (ROI) (FI_tumor_) and fluorescence intensity of tumor adjacent tissue ROI (FI_adjacent tissue_). For in vivo imaging, fluorescence intensities of six ROIs were selected from the gross HNSCC tumor area, and other six ROIs were selected from the surrounding non-tumorous tissues. Whenever possible, the ROIs were equally spaced and spread across each area in the surgical field. An overall mean fluorescence intensity (MFI) was calculated, using the mean fluorescence intensities from the ROIs. The MFI was defined as the total fluorescence signal divided by the total number of pixels within the ROI. The TBR for each tissue area was defined as the fluorescence signal of the tumor divided by the fluorescence signal of the surrounding normal tissue. Ex vivo imaging was used to determine the sensitivity and specificity of cMBP-ICG for surgical specimens. An ROC analysis was performed on data from the small tissue cut from specimens’ surfaces. The pathologist assessed whether the tumor was present in the tissue using a binary (yes/no) approach according to paraffin sections H&E stains.

Descriptive statistics, including the MFI and TBR, are presented as mean and SD. The MFI and TBR of the 10 in vivo lesions and ex vivo specimens from all of the patients were compared unpaired t test (two-tailed). We calculated the predictive values^17^ of the fluorescence signal of the fresh tissues against the H&E result. A P value of less than 0.05 was considered statistically significant. Statistics and figures were generated using GraphPad Prism (Version 8.0) and the R language *pROC* package (version 1.18.0)^18^ and *yardstick* package (version 0.0.9).

## 3. Results

### 3.1. cMBP-ICG Targets c-MET for Optical Imaging of Human HNSCC

To optimize imaging conditions, solutions of various concentrations (0–20 μM) of cMBP-ICG in ultrapure water were coated onto fresh specimens of porcine tongue. Pictures were captured from two different distances between object and lens at 20 and 40 cm. We found that the optimal conditions were the solution of either 2.5 or 5.0 μM and the imaging distance of 20 cm (**Figure S1**).

All patients (n=10) completed ex vivo fluorescence imaging, and 8 patients completed preoperative in vivo fluorescence imaging. In vivo imaging was not performed for patient 5 and patient 8 due to limitation of mouth opening. Patient 9 was a non-tumor patient, so it’s MFI was not included in TBR statistics. Both preoperative in vivo and intraoperative ex vivo imaging showed the pronounced fluorescence signal in the tumor area compared with the surrounding uninvolved mucosa for both coating concentrations. As the concentration doubled, the mean fluorescence intensity of both the tumor and the background increased. The preoperative imaging showed that the TBR of primary tumors was 4.12 ± 0.7 in the 5.0 μM group, and 4.08 ± 2.0 in the 2.5 μM group. The mean fluorescence intensity (MFI) of primary tumors and adjacent tissue was 65.09 ± 21.8 a.u. and 18.23 ± 8.3 in the 5.0 μM group, respectively, and 36.65 ± 30.9 a.u. and 10.21 ± 10.6 a.u. in the 2.5 μM group, respectively. The intraoperative imaging showed that the TBR of primary tumors was 2.71 ± 0.7 in the 5.0 μM group, and 3.11 ± 1.2 in the 2.5 μM group. The MFI of primary tumors and adjacent tissue was 47.35 ± 9.0 a.u. and 21.14 ± 8.2 in the 5 μM group, respectively, and 32.45 ± 34.2 a.u. and 13.38 ± 15.2 a.u. in the 2.5 μM group, respectively (**Figure 3**).

**Figure 3.**
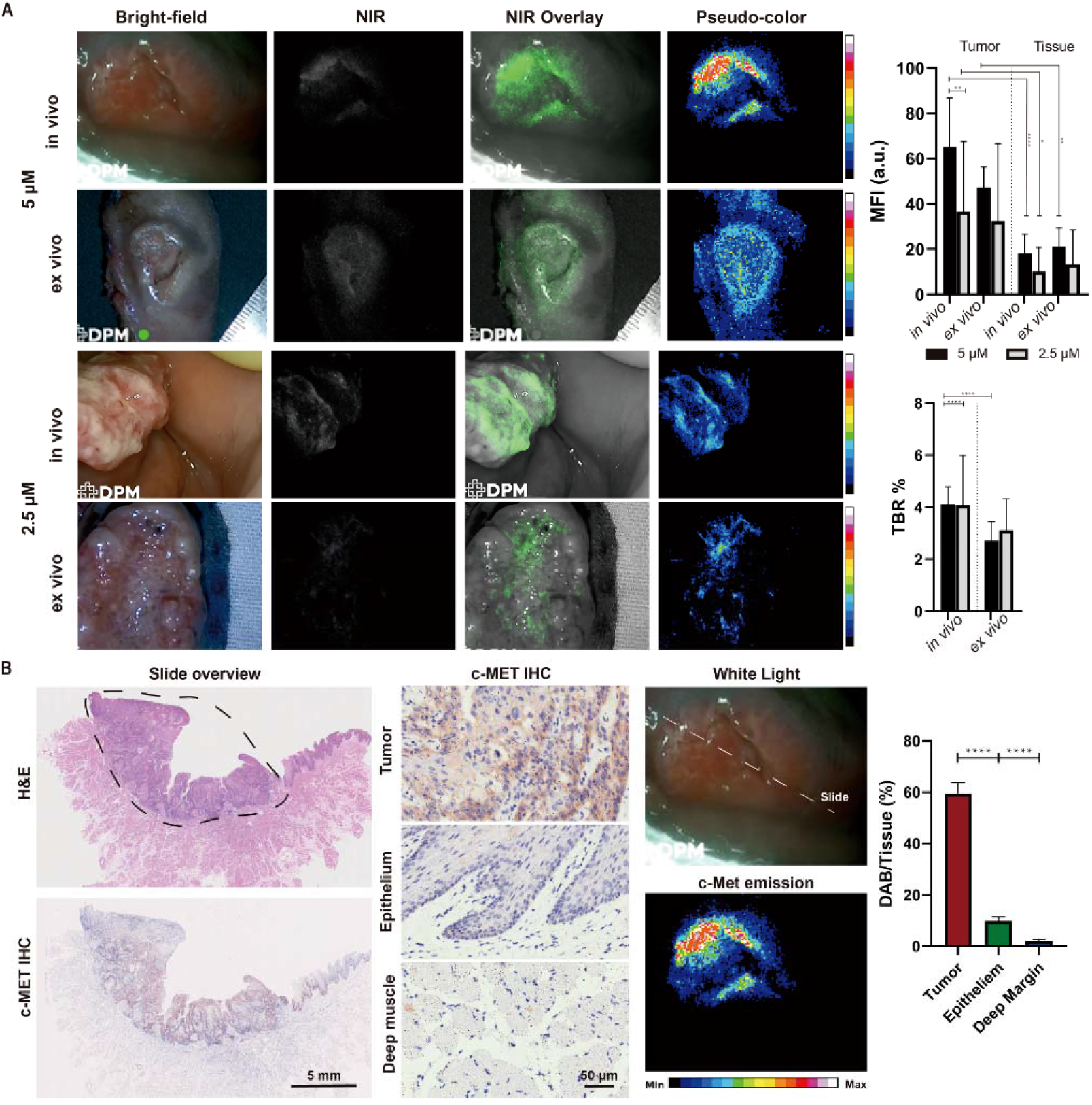
cMBP-ICG dosage selection, in*/*ex vivo analysis after fluorescence imaging, and correlation between c-MET expression and cMBP-ICG signal intensity. (A) The snapshots of two patients from different dosage groups. MFI and TBR of each group and each application procedure (in*/*ex vivo) were calculated following the same process. (B) Representative c-MET immunohistochemistry images and H&E images from a paraffin-embedded tumor section of a patient (slide overview), showing an area of HNSCC (black circle). Preoperative in vivo cMBP-ICG imaging of the patient shows the slide location in the whole tumor. *p < 0.05, **p < 0.01, ***p < 0.001, ****p < 0.0001. Data are presented as mean ± SEM.

Tissue samples from nine patients were available for c-MET expression analysis via IHC (excluding one non-tumor patient). c-MET IHC staining was analyzed and quantified in tumor, mucosal epithelium, and deep margin (i.e., healthy muscle tissue) based on the pathological assessment. The mean ratio of c-MET area (marked by DAB) over the total tumor tissue area was 57.3% ± 17.8%, which was significantly higher than that in normal mucosal epithelium (8.4% ± 3.6%) and in the deep muscular margin (4.1% ± 2.2%), indicating high c-MET expression in the tumor (**Figure 3B**, p < 0.0001 in both compared groups). The patients’ TBR, MFI, and DAB/Tissue area were summarized in **Figure S2**. To determine the specificity and sensitivity of cMBP-ICG for neoplastic tissue, the MFI data from the specimen cassettes was used to create an ROC curve. AUC, average positive predictive value (PPV), and negative predictive value (NPV) were showen in **Table 2**.

**Table 2.** Sensitivity and specificity of fluorescence for tumor surface samples

| samples | Group 1 | Group 2 | Both Groups | Surgeons |
| --- | --- | --- | --- | --- |
| Concentration | 5 μM | 2.5 μM | | |
| Sensitivity (%) | 100% | 46% | 71% | 60% |
| Specificity (%) | 75% | 88% | 83% | 95% |
| PPV | 80% | 79% | 83% | 93% |
| NPV | 100% | 61% | 73% | 71% |
| AUC | 0.88 | 0.65 | 0.78 | NA |
| 95% CI | 0.88-1 | 0.53-0.833 | 0.68-0.88 | NA |

### 3.2. cMBP-ICG Enables the Screening of Small Cancer Foci

A suspicious single lesion was detected 2 mm above the primary gum cancer area during preoperative in vivo NIR fluorescence imaging in one patient. The attending surgeon expanded the surgical removal area accordingly. This suspicious micro-lesion was visualized in intraoperative ex vivo NIR fluorescence imaging as well. An intraoperative frozen section was sliced with guidance from the ex vivo NIR fluorescence imaging, ensuring that the suspicious lesion was present in the frozen section. The pathology results verified this suspicious lesion as severe dysplasia of the epithelium, which had a high probability of cancerization **(Figure S3A**).

The attending surgeon suspected a gum leukoplakia in a patient to be a local metastasis considering its anatomic proximity to the primary tongue tumor. However, the NIR fluorescence intensity of this area in preoperative in vivo imaging was not higher than that of the adjacent normal tissue. This finding did not affect the operation plan. Still, the NIR fluorescence intensity of this area in the excised specimen was not higher than that in the adjacent normal tissue. The pathological analysis of frozen section of this area showed normal mucosa (**Figure S3B**).

### 3.3. cMBP-ICG for Rapid Biopsy and Tumor Grading

Biopsy in one patient revealed that one lesion of buccal mucosa was tumor-negative. The attending physician speculated that it was a tumor and that it needed to be removed. High intensity of both in vivo and ex vivo NIR fluorescence suggested partial cancerization (**Figure S4A**). Postoperative pathology results confirmed that the expression of c-MET in the high fluorescence signal area was high, and there were local small cancer foci in this area (**Figure S4B**).

In this study, the 10 cases included intraepithelial neoplasia (n = 1) (**Figure S5**A), well differentiated HNSCC (n = 2) (**Figure S5B**), moderately well differentiated HNSCC (low grade) (n = 6), and moderately differentiated HNSCC (intermediate grade) (n = 1) (**Figure S5**C). MFI and TBR of moderately well differentiated HNSCC and moderately differentiated HNSCC were analyzed by unpaired *t* test (**Figure S6**). There were significant differences in all outcomes. The degree of tumor differentiation greatly affected the 5-year survival rate of the patients. Our results suggests that cMBP-ICG can be used as an index of tumor differentiation degree, thereby improving treatment plan.

The topical usage of cMBP-ICG at a relatively high concentration did not result in any grade 1 or higher adverse events in patients with HNSCC.

## 4. Discussion

NIR fluorescence-guided surgery has emerged as a promising technique for head and neck cancers. The advantages of NIR ligands include increased penetration depth and lower autofluorescence compared with fluorophores emitting in the visible-wavelength range ^6,19-21^. Based on our previous work^22^, cMBP peptide was selected as the targeting ligand because of its high docking affinity to c-Met and known application in tumor therapy^9^ and imaging^10,23^. It’s dissociation constant (*K*_D_) is 3.96 × 10^−7^ M, as verified in our previous works^24^. This homing peptide may offer many properties including high tumor penetration, low immunogenicity, and cheap synthesis^12^. ICG was chosen as the imaging agent due to its biomedical safeness and high signal-to-background ratio^25^.

In this study, a coating concentration of 5.0 μM displayed better imaging contrast in a dark room during preoperative in vivo experiment. In contrast, under intraoperative ex vivo conditions, surgical lighting background in the operation theater caused higher background noise to sensors of the camera, and the concentration of 2.5 μM ensured a higher TBR. Regarding visual perception of real-time fluorescence imaging, the tumor area could be visualized more clearly in 5.0 μM group. High TBR from 2.5 μM group was contributed to low background noise, but it did not effectively help discriminate tumor regions. Therefore, MFI may be a more suitable index to referee tumor fluorescence imaging effectiveness. The ROC results in this study demonstrated that 5.0 μM group had higher AUC, PPV, and NPV, agreeing with the visual effect.

Because the ‘areal cancerization’ feature of HNSCC^26^, microscopic cancer foci or premalignant lesions may discretely occur around the primary cancer area. This characteristic is a key underling cause for the high relapse rate of HNSCC. We believe that the first patient mentioned in section 3.2 (whose occult lesion was detected by fluorescence imaging) benefitted from the approach used in this study. Our NIR fluorescence imaging method was proven capable in identifying and locating such small cancerous foci or precancerous lesions. The attending surgeon can revise surgical procedure accordingly to suppress the probability of cancer relapse.

HNSCC can invade various mucosa spaces through mucosa folds and tunnels. For the second patient mentioned in section 3.2, the suspicious leukoplakia was located in an area highly suggestive of a metastasis. Conservatively, surgeons are inclined to excide such lesions. However, oral environments of patients with oral cancer are usually unhealthy, and they exhibit mucocutaneous striatal abnormalities without pathological significance. Excessive excision may unnecessarily impair the physiological function. The NIR fluorescence imaging may assist the conventional technique in differentiating cancer foci for long-term follow-up of mucosal lesions and preventing overtreatment.

Biopsy is not very effective in detecting early-stage carcinoma in situ given that the cancerized area may be too small and biopsy puncture area may not cover the cancerized area. The aberrant activation and overexpression of c-MET is a very early event in the development of squamous cell carcinoma.^[42]^ Topical application of c-MET–targeting fluorescence dye was superior in detecting the early-stage cancer. c-MET expression varied with the severity of epithelial dysplasia. Specifically, the level of c-MET expression descended from carcinoma *in situ*, via highly atypical hyperplasia, to normal mucosa. For the patient mentioned in section 3.3, the observed NIR fluorescence intensity agreed well with this trend, reflecting c-MET expression. Therefore, topical application of cMBP-ICG can be used to guide biopsy sampling, and may shift the detection of HNSCC to an earlier time point.

The study has many strengths. First, topically applied real-time imaging agents are well-suited for the study of head and neck cancer^11^. Given that HNSCC develops from the mucosal epithelium in the oral cavity, pharynx, and larynx^27^, and is present on the oral and oropharyngeal surfaces, gargling may be an excellent choice for noninvasive and convenient prompt investigation. Namely, the mouth-washing topical application of the targeting imaging agents may be an optimal approach given that mouthwash covers the entire oral mucosa and directly screens out suspicious small cancer foci without restriction from the other tissues. Secondly, being applied topically, the imaging time of NIR fluorescence probes would be significantly reduced (within minutes)^11,14,28^. Peptide-based targeting agents’ fast renal clearance^12^ is not seen as a drawback, but rather a benefit from the perspective of medication safety.

Relatively small sample size limited this research. The tumor tissues resided sub-surface. NIR fluorescence imaging was conducted surfaces of ROI, lacking imaging information of cross-section of tumor tissues. Relevant study regarding in-depth imaging is ongoing. NIR imaging obtained in this study cannot be compared with the ones obtained via IV injection.

## 5. Conclusion

In conclusion, the topical application of cMBP-ICG shows good biosafety and improved selectivity for HNSCC compared with local injection. Our clinical trial demonstrated the remarkable ability of cMBP-ICG to mark tumor margins, reveal concealed minor tumor foci, visualize lymph nodes metastases with ENE, and index HNSCC differentiation grades. One patient benefited from this study, i.e., surgery excision boundaries were expanded due to the results of NIR fluorescence imaging. Our results provide evidence that topical application of c-MET–targeting fluorescence probe is a quick, easy, and economical method to detect HNSCC. c-MBP-ICG has a potential in guiding biopsy and surgical procedure, long-term follow-up of leukoplakia, and postoperative evaluation of HNSCC patients.

## Data Availability

All data produced in the present study are available upon reasonable request to the authors.

## Acknowledgements

Jingbo Wang, Siyi Li, and Kun Wang contributed equally to this work.

This work was supported by funds from the The National Natural Science Foundation of China (nos. 91859202 and 82172049).

The authors wish to acknowledge Dr Changyou Zhan, professor of Fudan University, for insightful discussion.

