## Supplemental Information for "Surgical Guidance of Head and Neck Squamous Cell Carcinoma Enabled by a c-MET-Targeting Fluorescent Probe cMBP-ICG"

**Abstract:**

Surgical procedure guided by near infrared (NIR) fluorescence dye, that is administrated via intravenous injection, is appealing to researchers and doctors. Tumors such as head and neck squamous cell carcinoma (HNSCC), which is shallow, invasive, and heterogenous, hampered such effort due to poor delivery. Herein, we applied a peptide-based NIR fluorescence dye, cMBP-ICG, which can be administrated via topical application and specifically target tumor tissue via c-MET binding, in a clinical trial. Ten clinical cases, which were diagnoses (suspectable) HNSCC by attending physician with biopsy, were included. cMBP-ICG solutions were applied with gargle onto tissue surface *in vivo* preoperatively. One case with non-tumor lesion, one case with biopsy-negative carcinoma *in situ*, and one case with additional foci (excision area were extended according), were screened out. Intraoperative *ex vivo* fluorescence imaging, in which cMBP-ICG was coated onto mucosa surfaces and cross-sections of specimens, clearly outlined horizontal and vertical margins of the tumor. Its efficiency was not diminished by interference of previous chemo- and radio-therapy. Twenty lymph nodes were also imaged upon cMBP-ICG coating, in which two out of two tumor-positive nodes were screened out with an accuracy of 100%. Fluorescence intensity from cMBP-ICG was found positively corelated to differentiation degree of HNSCC. In summary, topical application of cMBP-ICG was capable of differentiating preoperative tumor, revealing concealed foci, outlining margin of tumor intraoperatively, and detecting metastasized lymph nodes.

**Key words：**

near infrared fluorescence dye, topical application, c-MET, head and neck squamous cell carcinoma

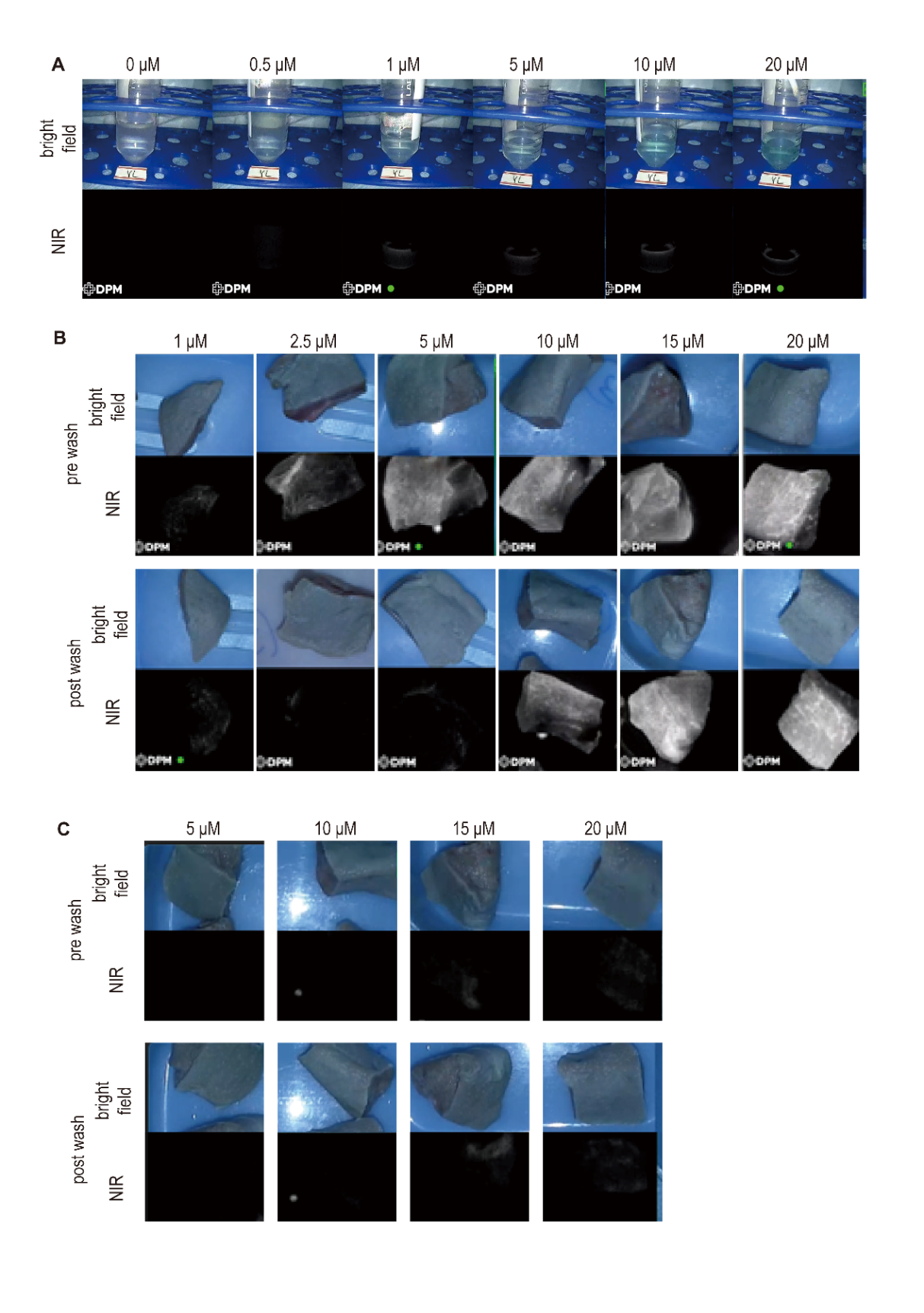

**Figure S1.** Evaluation of sensitivity of the imaging system using cMBP-ICG containing (A) ultrapure water phantoms (0–20 μM). (B, C) Fresh specimens of porcine tongue were coated with cMBP-ICG, followed by a cleansing procedure. Pictures were taken as references to identify the best dosage for topical application. Pictures taken from variable distance (B. 20 cm vs. C. 40 cm).

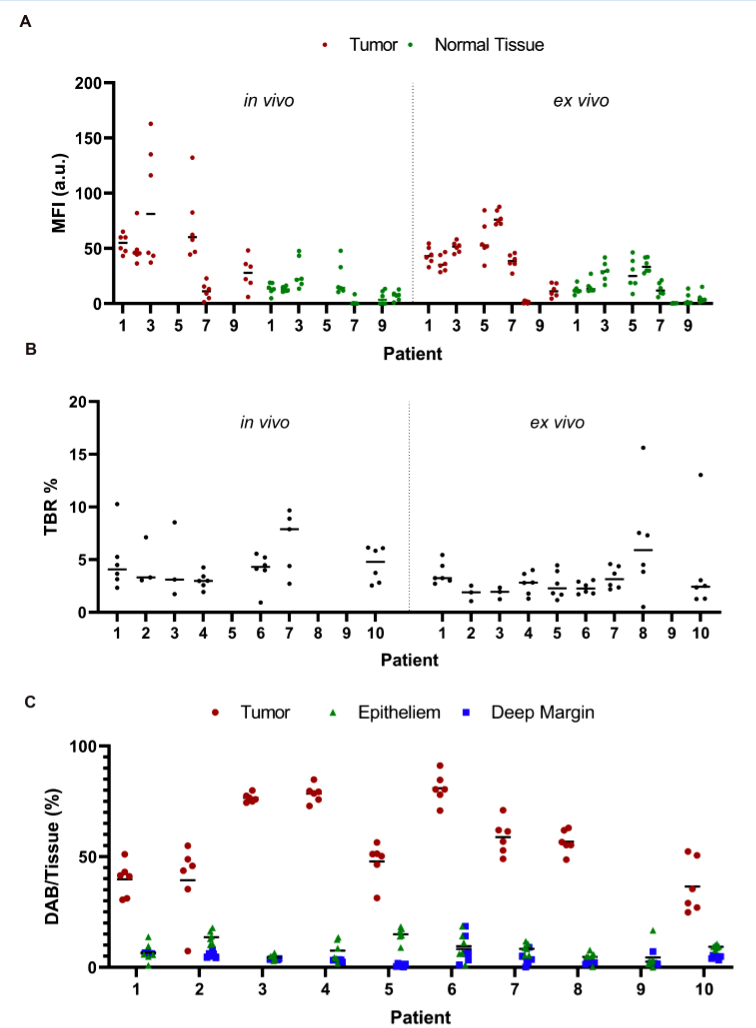

**Figure S2**. Quantification of c-MET expression in immunohistochemistry, immunofluorescence samples, and pre-/intraoperative cMBP-ICG florescence (n = 10 patients). MFI (A), TBR (B), and DAB/Tissue area (C) were measured with Pearson correlation. In vivo imaging was not performed for patient 5 and patient 8 due to limitation of mouth opening. Patient 9 was a non-tumor patient, so it’s MFI was not included in TBR statistics.

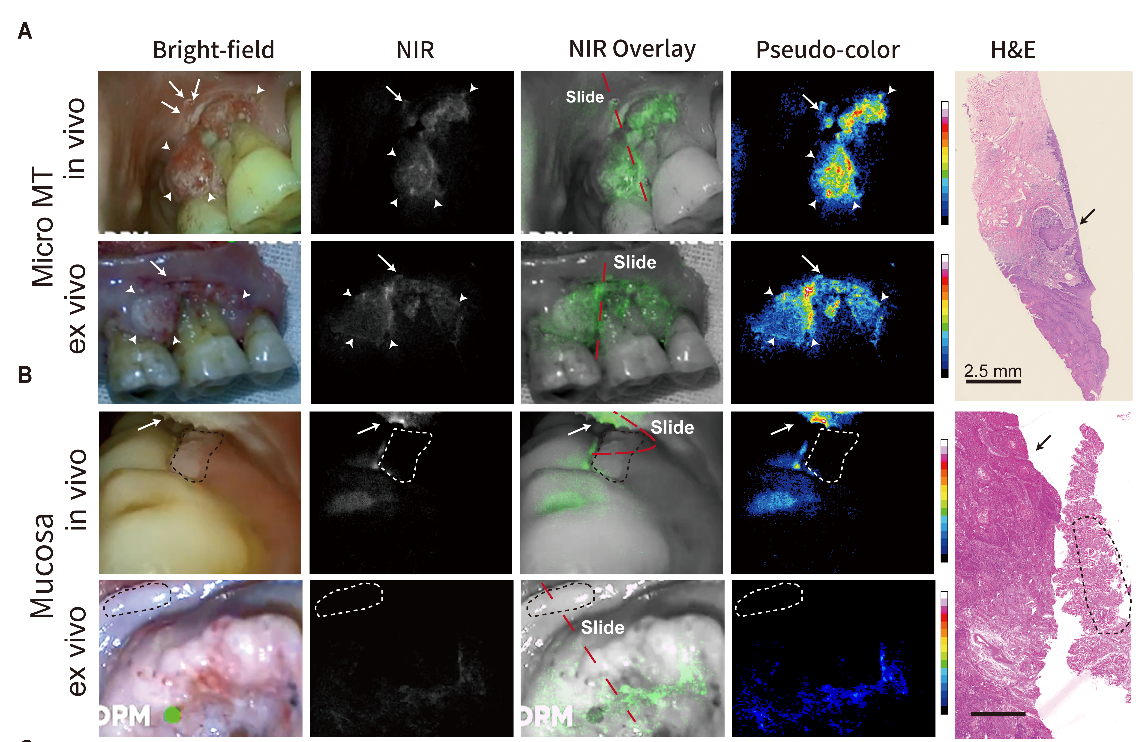

**Figure S3**. Suspicious lesions discovered (A), and a nontumor lesion screened out (B) during preoperative imaging procedure and lymph nodes with or without metastasis (C). Frozen sections were cut according to the real-time videos. The section locations (red dashed line) were ensured to meet the region of interest. (A) Severe dysplasia (arrow) was detected 2 mm above the primary gum cancer area (arrowhead). (B). A gum leukoplakia (dashed circle) adjacent to primary tongue cancer (arrow) was proved to be normal mucosa pathologically. White arrow = cancer foci; white dashed circle = suspicious lesion. MT = malignant tumor; NIR = near infrared; H&E = hematoxylin and eosin.

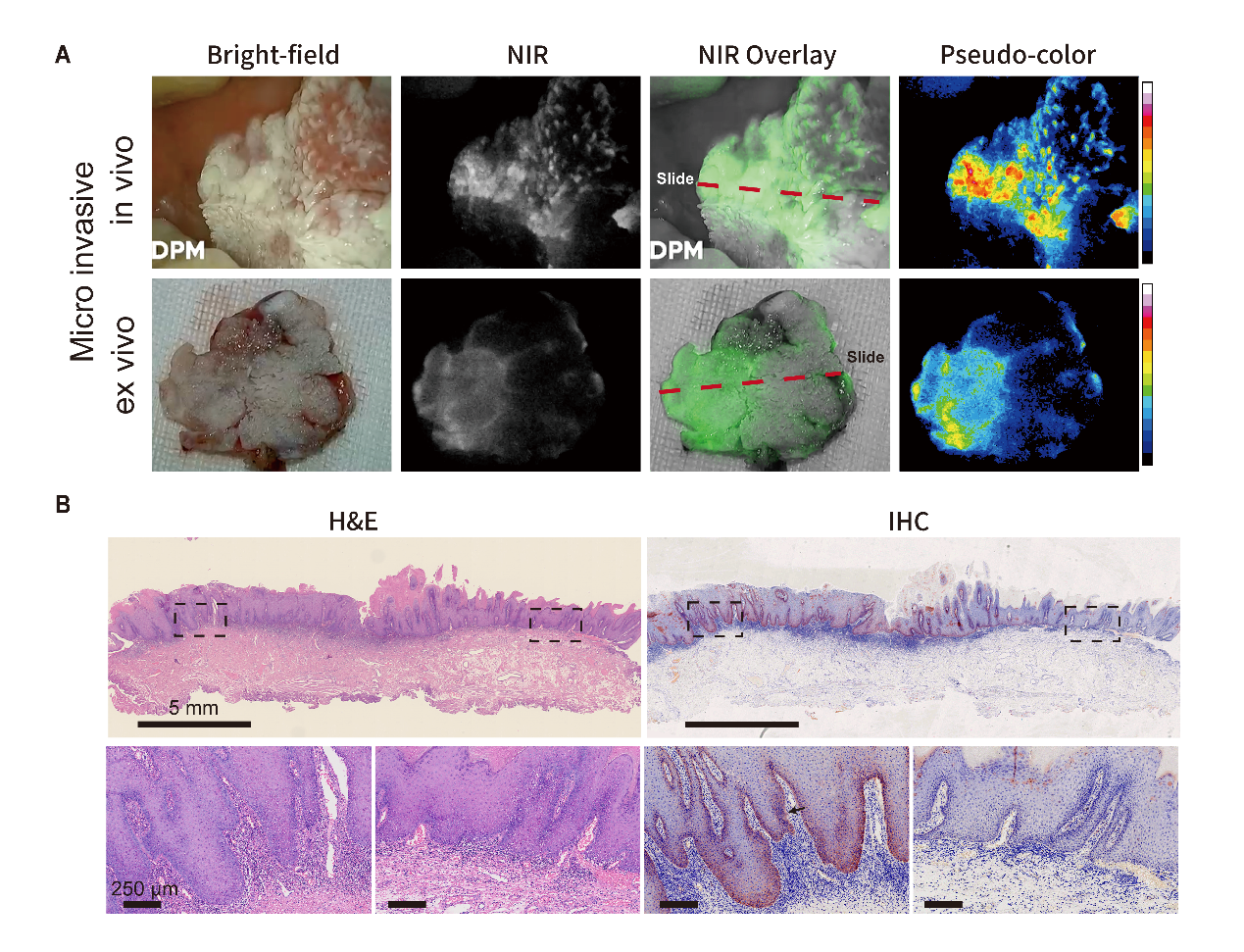

**Figure S4.** Heavily keratinized mucosa with small cancer foci in one patient. (A) The snapshots from preoperative (upper row) and intraoperative (lower row) imaging. (B) H&E and IHC of the frozen sections. The lower row demonstrates enlarged views of the two areas framed in black in the upper row. The section locations (red dashed line in panel A) were ensured to meet the region of interest. H&E = hematoxylin and eosin; IHC = immunohistochemistry.

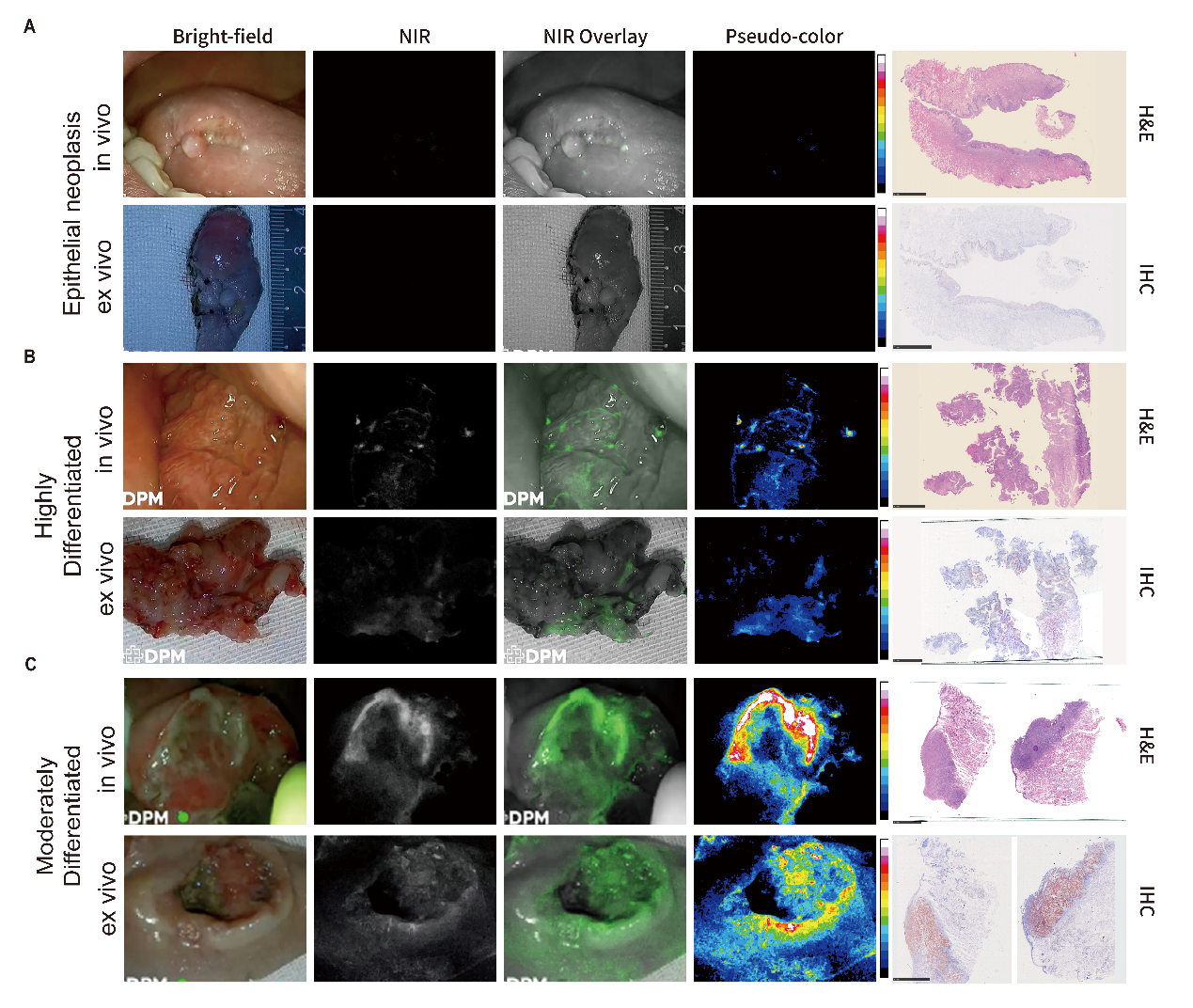

**Figure S5.** Comparison of epithelial neoplasia and two grades of HNSCC. (A) Epithelial neoplasia showed low FI in both in/ex vivo imaging. The frozen section showed low c-MET expression of this lesion. (B) Highly differentiated HNSCC showed slightly increased FI compared with panel A. The frozen section showed about 25% of c-MET expression area fraction. (C) Moderately differentiated HNSCC showed high FI in in*/*ex vivo imaging. The frozen section showed about 90% of c-MET expression area fraction. FI = fluorescence intensity.

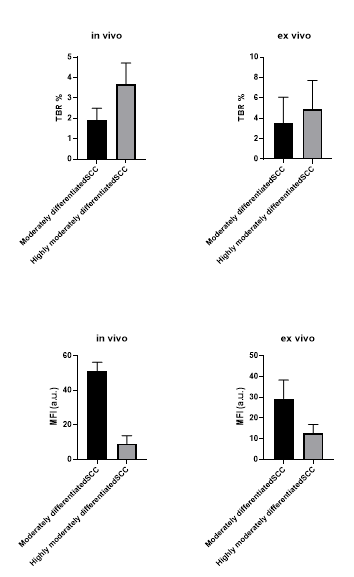

**Supplementary Figure S7**. Unpaired *t* test for TBR and MFI of highly moderately differentiated and moderately differentiated HNSCC.
